# A Vision-Language Model for Coronary Angiography Interpretation and Clinical Decision Support

**DOI:** 10.64898/2026.08.11.26360095

**Authors:** Zhixing Li, Yixuan Sun, Chen Jiang, Tan Pan, You Zhou, Cong Wang, Lei Pan, Xingmeng Zhang, Zhenqi Yang, Ziqing Yu, Zilong Xiao, Juecheng Chen, Yunshi Huang, Rui Sun, Yunhui Gan, Xiao Li, Beijian Zhang, Zhi Zhang, Xiansheng Wang, Limei Han, Yuan Qi, Shuo Wang, Yuan Cheng, Yixiu Liang, Junbo Ge

## Abstract

**BACKGROUND:** Coronary angiography remains the reference standard for diagnosing coronary artery disease and guiding revascularization, yet its interpretation requires expert integration of multi-view anatomy, lesion morphology and procedural context. Existing artificial intelligence approaches are largely task-specific, annotation-dependent and limited in capturing the semantic relationship between angiographic findings and interventional decision-making. Whether large-scale vision-language pretraining can enable transferable foundation-model representations for invasive coronary imaging remains unknown.

**METHODS:** We developed CAG-MIND, a domain-specific vision-language foundation model for coronary angiography, using 135,475 paired coronary angiography (CAG)–procedural report cases comprising 812,850 angiographic videos collected from Zhongshan Hospital and Shanghai Geriatric Medical Center. Each case consisted of standardized six-view angiographic acquisitions paired with structured procedural semantics extracted from routine reports using a large language model-assisted pipeline. The model was pretrained by aligning multi-view angiographic representations with report-derived semantic embeddings through bidirectional contrastive learning. Performance was evaluated under zero-shot and supervised fine-tuning settings across 11 downstream tasks grouped into structural abnormality detection, atherosclerotic plaque assessment, and interventional decision prediction, using both an internal validation cohort and an independent external test cohort.

**RESULTS:** CAG-MIND demonstrated robust performance across all three task categories. In the zero-shot setting, the model achieved mean AUROCs of 0.686 in the internal validation cohort and 0.745 in the external test cohort, indicating transferable multimodal representations without task-specific supervision. Following supervised fine-tuning, the mean AUROC increased to 0.827 and 0.846, respectively, with excellent performance for coronary stenosis detection (AUROC 0.940 in both cohorts), balloon/stent prediction (0.900 and 0.907), and CABG recommendation (0.877 and 0.875). Compared with representative biomedical vision-language models and conventional image-based architectures, CAG-MIND consistently achieved superior performance in both zero-shot and supervised settings and remained superior to fully fine-tuned competing models when trained with only 10% of the labelled data. Grad-CAM visualization demonstrated anatomically plausible lesion-focused attention, supporting the interpretability of the learned representations.

**CONCLUSIONS:** CAG-MIND is, to our knowledge, the first large-scale vision-language foundation model for coronary angiography trained at more than 100,000-patient scale. By aligning standardized multi-view angiographic videos with report-derived procedural semantics, CAG-MIND enables robust zero-shot transfer, data-efficient fine-tuning and cross-center generalization. These findings support domain-aligned multimodal pretraining as a scalable foundation-model paradigm for invasive cardiovascular imaging and future cath-lab decision support.

## Introduction

Coronary angiography (CAG) remains the clinical gold standard for the diagnosis and management of coronary artery disease (CAD), guiding both diagnostic evaluation and interventional treatment planning [1,2]. Clinical interpretation of CAG requires comprehensive integration of coronary anatomy, lesion morphology, vessel distribution, haemodynamic relevance, and procedural feasibility across multiple angiographic projections, making image interpretation highly dependent on operator expertise and clinical experience [3,4]. Although modern catheterization laboratories generate vast quantities of coronary angiographic videos accompanied by routine procedural reports, these multimodal clinical data are primarily used for individual patient care and remain largely underexploited as a scalable resource for learning generalizable representations of coronary disease.

Recent advances in artificial intelligence (AI) have enabled automated analysis of cardiovascular imaging across multiple modalities, including echocardiography [5,6] and electrocardiography [7,8]. In coronary CT (computed tomography) angiography, AI has been applied to a range of clinically relevant tasks, including stenosis detection, quantitative lesion assessment and plaque characterization, and procedural outcome prediction [9–11]. These approaches have demonstrated the potential of AI to improve efficiency, consistency, and decision support during coronary intervention [12]. However, they are typically optimized using manually curated labels for predefined tasks, limiting their ability to learn transferable representations that generalize across the broader diagnostic and interventional workflow. As a result, most existing approaches require independent model development for each downstream application and fail to capture the hierarchical clinical relationships linking coronary anatomy, plaque characteristics, and treatment decisions.

Vision-language foundation models (VLMs) have recently emerged as a powerful paradigm for scalable multimodal representation learning by aligning imaging data with routinely generated clinical text through natural language supervision [13–16]. Rather than depending on exhaustive manual annotation, these models leverage routine clinical documentation as weak supervision to capture rich semantic information embedded within real-world clinical workflows, enabling transferable representations that generalize across diverse downstream tasks. Cardiovascular imaging studies, including echocardiography-based multimodal foundation models, have further demonstrated the feasibility of learning clinically meaningful representations directly from paired imaging and clinical reports [17,18]. These advances suggest that vision-language foundation models provide a promising framework for overcoming the limitations of conventional task-specific AI systems and establishing more generalizable AI models for cardiovascular imaging.

Here, we sought to develop CAG-MIND, a large-scale vision-language foundation model for coronary angiography by leveraging routinely acquired angiographic videos paired with corresponding procedural reports. We further aimed to establish a standardized multimodal learning framework capable of learning transferable representations of coronary anatomy and procedural semantics, thereby providing a unified foundation for diverse downstream diagnostic and interventional applications in invasive cardiovascular imaging.

## Methods

### 1. Study Design and Data Collection

We conducted a retrospective, multi-center study to develop and evaluate CAG-MIND (Coronary AngioGraphy Multimodal INtelligence for Diagnosis/Decision support), a domain-specific vision-language foundation model for coronary angiography. Consecutive invasive coronary angiography examinations were collected from Zhongshan Hospital, Fudan University, and Shanghai Geriatric Medical Center. Each case consisted of coronary angiographic cine videos and the corresponding procedural documentation, including angiographic findings, procedural records and clinical conclusions.

After patient-level deduplication, longitudinal record reconciliation, file-integrity screening and quality control, the final dataset comprised 135,475 patients and 812,850 angiographic videos. Each patient-level case was represented by six standardized angiographic views to ensure anatomically comprehensive visualization of the coronary tree and to reduce heterogeneity caused by variable acquisition protocols. Cases with incomplete video data, missing procedural reports, corrupted files, failed projection verification, or insufficient multi-view coverage were excluded.

The dataset was divided into an internal training cohort, an internal validation cohort and an independent external test cohort. The internal cohorts were derived from Zhongshan Hospital and included 107,839 training cases and 23,454 validation cases. The external test cohort included 4,182 cases from Shanghai Geriatric Medical Center and was used to assess cross-center generalizability under institutional distribution shift.

**Figure 1.**
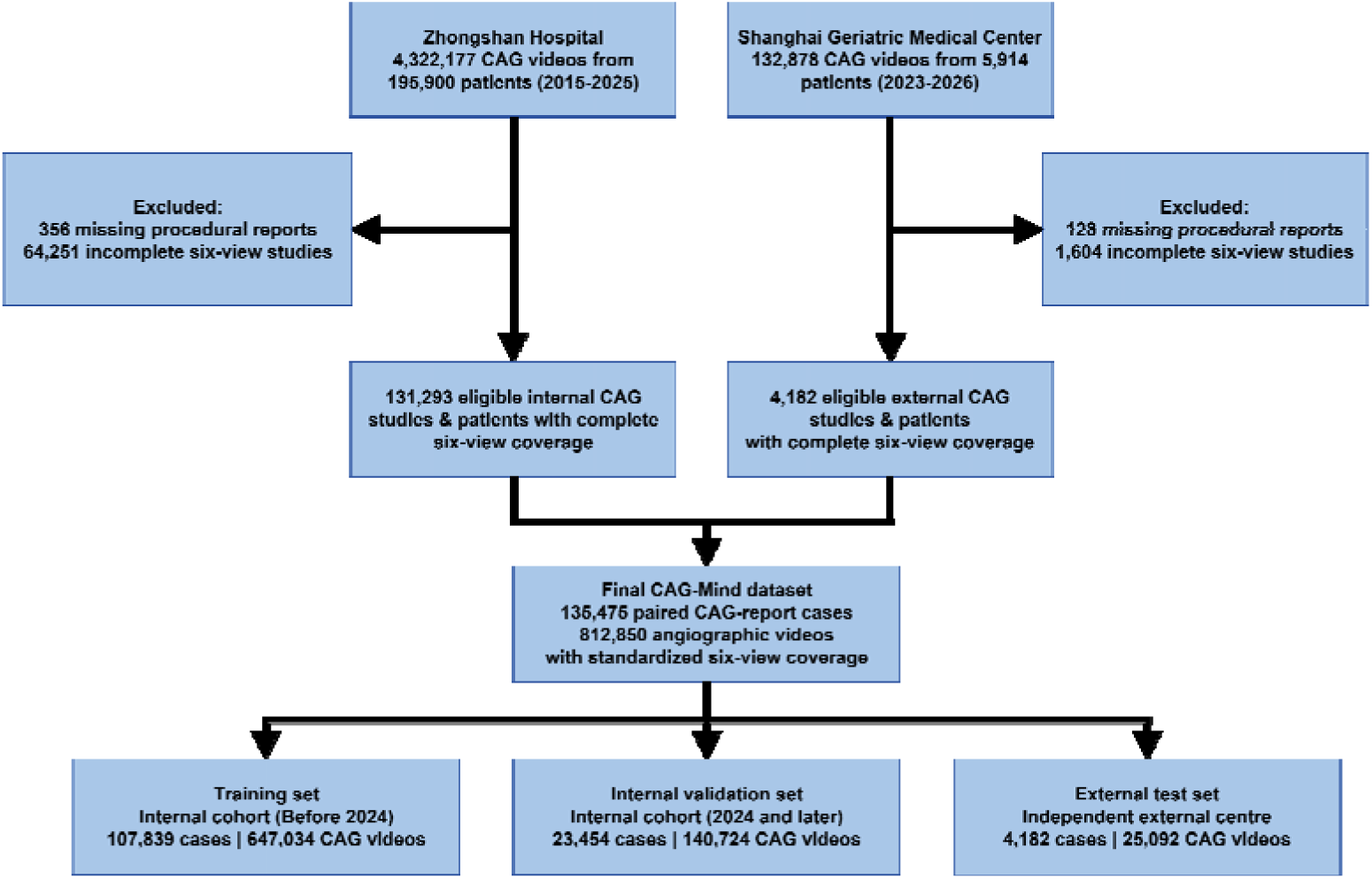
Construction of the CAG-MIND dataset and study cohorts. Workflow illustrating dataset assembly, eligibility screening, and cohort allocation for model development and evaluation. Coronary angiography (CAG) studies were collected from Zhongshan Hospital, Fudan University (2015–2025) and the Shanghai Geriatric Medical Center (2023–2026). Studies with missing procedural reports or incomplete standardized six-view angiographic coverage were excluded. The final CAG-MIND dataset comprised 135,475 paired coronary angiography–procedural report cases (812,850 angiographic videos). The Zhongshan Hospital cohort was chronologically divided into an internal training set (before 2024) and an internal validation set (2024 onward), whereas the Shanghai Geriatric Medical Center cohort served as an independent external test set. CAG, coronary angiography.

### 2. Angiographic Video Processing and Anatomical Standardization

All angiographic videos were processed through a dedicated curation pipeline that included data format harmonization, video integrity checking, patient-level matching, projection verification and standardized view assignment. Because coronary angiography interpretation depends on complementary projections rather than a single view, each case was organized into a six-view representation framework. This framework was designed to capture left and right coronary anatomy across clinically relevant projection angles and to provide consistent anatomical coverage across institutions.

For each case, the six angiographic videos were treated as a unified patient-level imaging input. View-level features were extracted independently and then fused to generate a global multi-view representation of coronary anatomy, lesion morphology and procedure-relevant visual context. This design allowed the model to integrate complementary information across projections while preserving view-specific angiographic features.

**Fig. 2.**
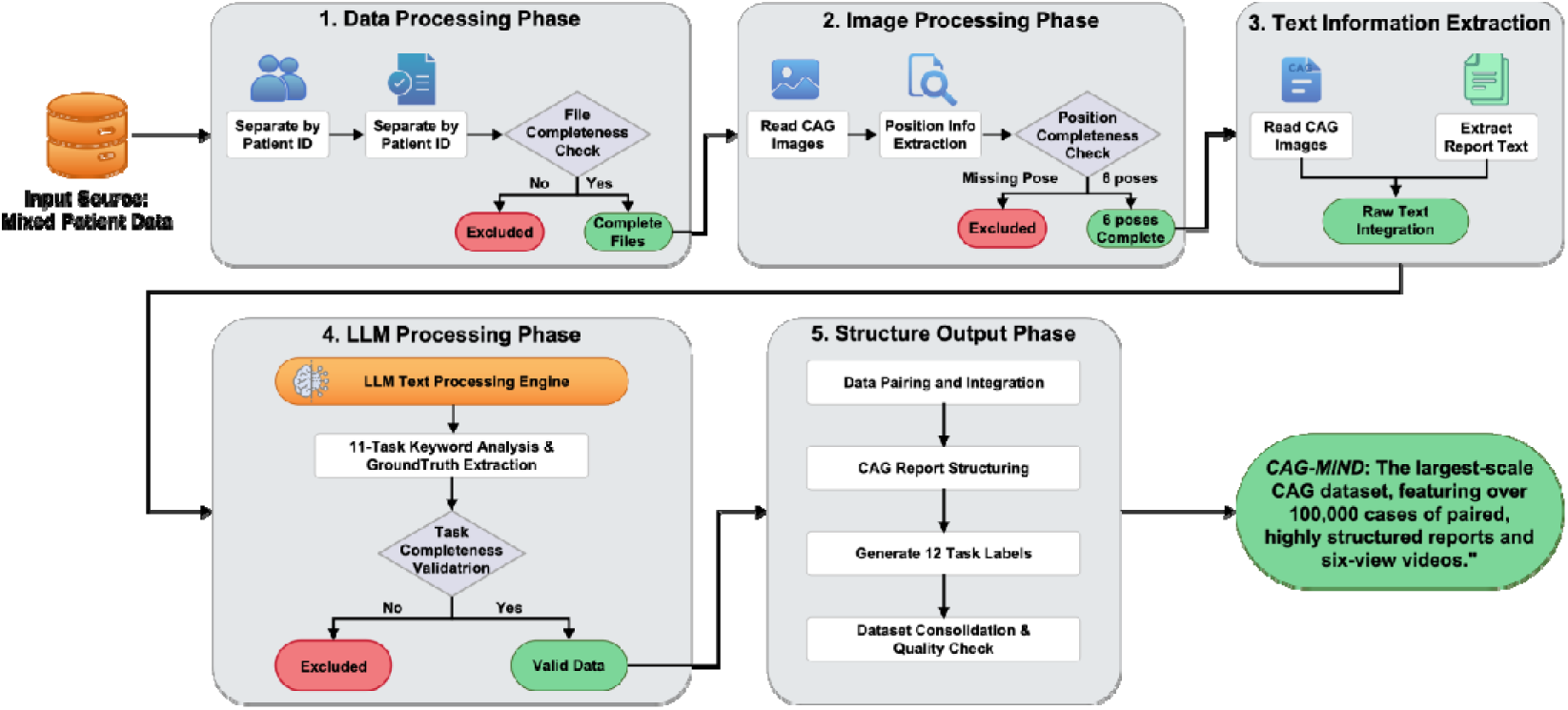
Overview of the CAG-MIND dataset construction pipeline. Paired coronary angiography (CAG) videos and reports were processed through five stages -- data processing, image processing, text information extraction, LLM-based text processing and structured output generation -- to construct a large-scale dataset comprising more than 100,000 paired cases, with structured reports, task labels and quality control.

### 3. Report Processing and Structured Semantic Extraction

Procedural reports were collected in raw textual form and exhibited heterogeneity in formatting, terminology and reporting style. To convert routine clinical documentation into scalable computational supervision, we developed a large language model-assisted semantic extraction pipeline. Reports were first decoded and standardized across formatting conventions. Task-specific information was then extracted and mapped to predefined diagnostic and therapeutic endpoints.

The semantic extraction framework generated structured labels for 11 clinically relevant endpoints, including coronary stenosis, calcification, prior stent, coronary ectasia, plaque rupture, thrombus, balloon or stent requirement, coronary artery bypass grafting recommendation, rotational atherectomy requirement, thrombus aspiration and intravascular imaging use. These endpoints were selected to cover three major domains of coronary angiography interpretation: structural abnormality detection, atherosclerotic plaque assessment and interventional decision prediction.

The language model-based extraction process included numerical value recognition, entity parsing, cross-sentence reasoning and logical consistency verification. Automated validation rules were used to identify internally inconsistent outputs, such as incompatibility between lesion severity descriptions and procedural decisions. Cases that failed completeness or consistency checks were either corrected through secondary parsing when possible or excluded from the final structured dataset. The resulting report-derived labels provided scalable weak supervision without requiring exhaustive manual annotation.

### 4. Model Development

CAG-MIND was developed as a vision-language foundation model that aligns multi-view angiographic representations with structured procedural semantics. The framework consisted of four major components: a report semantic encoder, a multi-view coronary angiography encoder and fusion module, a cross-modal memory module and a bidirectional contrastive learning module. The report semantic encoder transformed structured procedural information into semantic embeddings representing anatomical findings, plaque characteristics and procedure-related decisions. In parallel, the angiographic encoder extracted view-specific visual features from the six standardized cine projections. These view-level features were integrated through a multi-view fusion module to form a unified patient-level representation of coronary anatomy and lesion appearance.

To stabilize cross-modal representation learning at scale, angiographic and report embeddings were maintained in modality-specific memory banks. These memory banks provided cross-batch reference features and improved the robustness of contrastive alignment. The visual and textual representations were then projected into a shared latent space and optimized using bidirectional contrastive learning. The model was trained to maximize the similarity between paired angiographic videos and their corresponding report-derived semantic representations, while minimizing similarity to non-matched cases within the batch and memory banks.

This design explicitly linked angiographic morphology with structured clinical interpretation and interventional intent. Unlike conventional supervised classifiers trained separately for individual endpoints, CAG-MIND was trained to learn a generalizable multimodal representation of coronary angiography that could be transferred to multiple downstream diagnostic and therapeutic tasks.

**Fig. 3.**
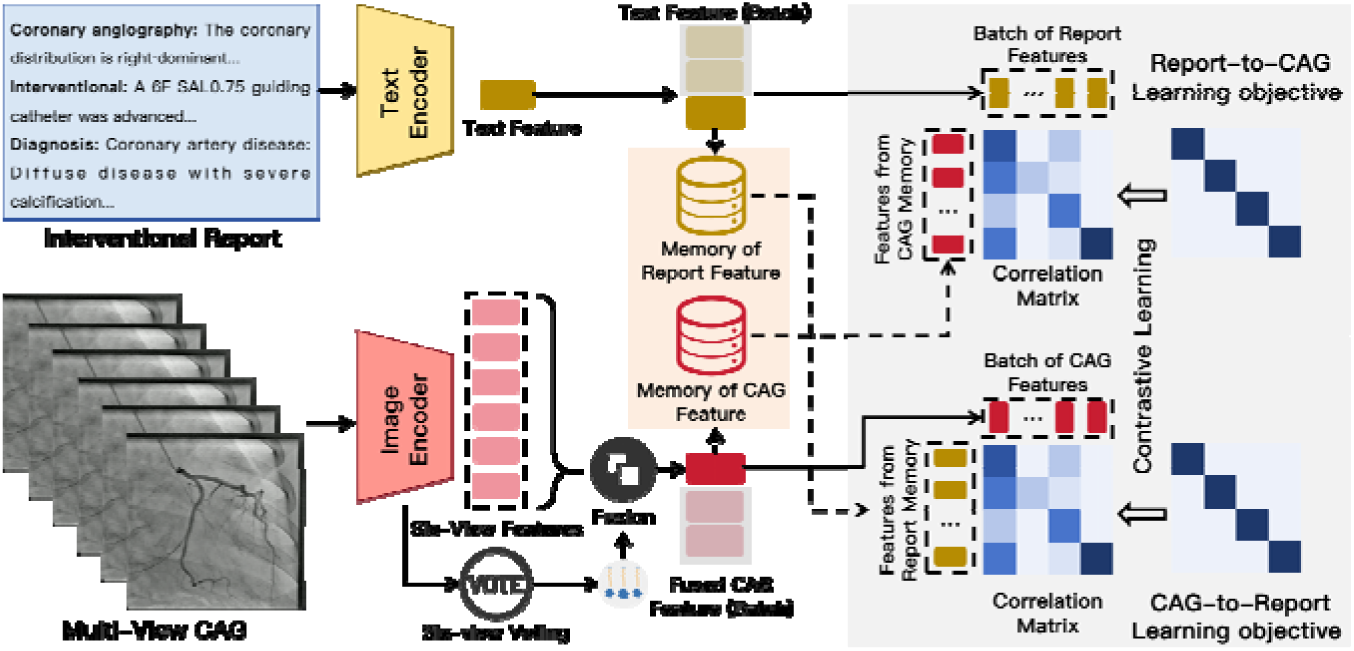
Overview of the cross-modal learning framework. The framework comprises four components: a report semantic encoder, a multi-view coronary angiography (CAG) fusion module, a cross-modal memory module and a bidirectional contrastive learning module. Structured reports and multi-view CAG videos are encoded into modality-specific representations, which are stored in separate memory banks and aligned through bidirectional contrastive learning.

### 5. Model Evaluation

Model performance was evaluated in both zero-shot and supervised fine-tuning settings. In the zero-shot setting, the pretrained vision-language model was evaluated without task-specific supervised optimization. This analysis was designed to assess whether multimodal pretraining alone could generate transferable representations for clinically relevant coronary angiography tasks. For each endpoint, task-specific textual prompts or structured semantic prototypes were compared with angiographic representations in the shared embedding space to generate prediction scores.

In the supervised setting, the pretrained visual representation was fine-tuned using labelled cases from the internal training cohort. Fine-tuning was performed for all 11 endpoints, and performance was evaluated in both the internal validation cohort and the independent external test cohort. To assess data efficiency, an additional experiment fine-tuned CAG-MIND using only 10% of the internal training data. This experiment was used to determine whether domain-aligned pretraining could reduce the amount of task-specific labelled data required for downstream adaptation.

The primary performance metric was the area under the receiver operating characteristic curve (AUROC). Task-level AUROCs were calculated for each diagnostic and therapeutic endpoint, and mean AUROC across all 11 tasks was used to summarize overall performance. Internal validation assessed in-distribution performance, whereas external testing assessed model robustness under cross-center domain shift.

### 6. Model Comparison

CAG-MIND was compared with representative vision-language and image-based baseline models. In zero-shot evaluation, CAG-MIND was compared with BiomedCLIP, a general biomedical vision-language foundation model, and CLIP-CAG, a coronary angiography-adapted CLIP-style baseline. This comparison was designed to evaluate whether coronary angiography-specific multimodal pretraining provides advantages beyond general biomedical image-text representation learning and conventional CLIP-style adaptation.8

In supervised fine-tuning experiments, CAG-MIND was compared with 3DCNN, Xception, Swin3D and fine-tuned CLIP-CAG. These baselines represented conventional convolutional neural networks, video-based deep learning architectures, transformer-based visual models and vision-language models adapted to coronary angiography. All models were trained or fine-tuned using the same internal training cohort and evaluated on the same internal validation and external test cohorts. Model comparisons were performed across the identical 11 endpoints to ensure task-level consistency.

The comparison strategy was designed to address three questions: whether CAG-MIND improves zero-shot transfer, whether its pretrained representation provides a stronger initialization for supervised fine-tuning, and whether its performance advantage persists under external validation.

### 7. Model Explainability

To assess whether CAG-MIND relied on anatomically and pathologically plausible image regions, we used Gradient-weighted Class Activation Mapping (Grad-CAM) to visualize spatial attention patterns in representative angiographic cases. Grad-CAM maps were generated for selected diagnostic tasks, including coronary stenosis and coronary ectasia, using the fine-tuned model.

The activation maps were overlaid on the corresponding angiographic frames to evaluate whether prediction-relevant regions were localized to coronary vessel segments, lesion-bearing areas or irrelevant background structures. View-specific activation patterns were also examined to determine whether the model adaptively used complementary information from different angiographic projections. These analyses were intended to provide qualitative evidence of anatomical plausibility rather than definitive proof of causal reasoning.

### 8. Statistical Analysis

Performance was summarized using AUROC for each endpoint and mean AUROC across all 11 tasks. Results were reported separately for the internal validation cohort and the independent external test cohort. Zero-shot and fine-tuned performance were compared across CAG-MIND and baseline models. Cross-center robustness was assessed by comparing performance patterns between the internal and external cohorts. Class imbalance was preserved in the evaluation cohorts to reflect real-world disease prevalence and procedural practice patterns.

## Results

### 1. Patient characteristics

A total of 135,475 patients were included in this study, comprising 107,839 patients in the internal training cohort, 23,454 in the internal validation cohort, and 4,182 in the external test cohort. The demographic characteristics were broadly comparable across the three cohorts. The mean age was 63.24 ± 10.01 years in the training cohort, 63.57 ± 10.47 years in the validation cohort, and 64.34 ± 10.43 years in the external test cohort, with median ages of 64 years (IQR, 57–70), 65 years (IQR, 57–71), and 66 years (IQR, 58–72), respectively. Male patients accounted for 70.8%, 74.0%, and 69.5% of the training, validation, and external cohorts, respectively, indicating a predominance of men across all datasets (Table 1).

**Table 1.** Baseline characteristics of the study population across the internal training, internal validation, and external test cohorts. Data are presented as *n* (%), mean (standard deviation [s.d.]), median (interquartile range [IQR]), or range, as appropriate. The internal training and validation cohorts were derived from Zhongshan Hospital, Fudan University, and the external test cohort was obtained from Shanghai Geriatric Medical Center. IQR, interquartile range; s.d., standard deviation.

| Characteristic | Internal training | Internal validation | External test |
| --- | --- | --- | --- |
| **n** | 107,839 | 23,454 | 4,182 |
| **Age, mean (s.d.)** | 63.24 (10.01) | 63.57 (10.47) | 64.34 (10.43) |
| **Age, median (IQR)** | 64.0 (57.0–70.0) | 65.0 (57.0–71.0) | 66.0 (58.0–72.0) |
| **Age, range** | 16–98 | 16–97 | 27–97 |
| **Male, n (%)** | 76,365 (70.8) | 17,360 (74.0) | 2,908 (69.5) |
| **Female, n (%)** | 31,474 (29.2) | 6,094 (26.0) | 1,274 (30.5) |

### 2. Zero-shot performance across coronary angiography tasks

We first evaluated the zero-shot performance of CAG-MIND across 11 clinically relevant coronary angiography tasks, which were organized into three hierarchical categories: structural abnormality detection (coronary stenosis, calcification, prior stent, and coronary ectasia), atherosclerotic plaque assessment (plaque rupture and thrombus), and interventional decision prediction (balloon/stent requirement, CABG recommendation, rotational atherectomy, thrombus aspiration, and intravascular imaging) (Fig. 4a,b and Table 2). Zero-shot evaluation was performed without task-specific supervision to assess the intrinsic transferability of the learned multimodal representations.

**Figure 4.**
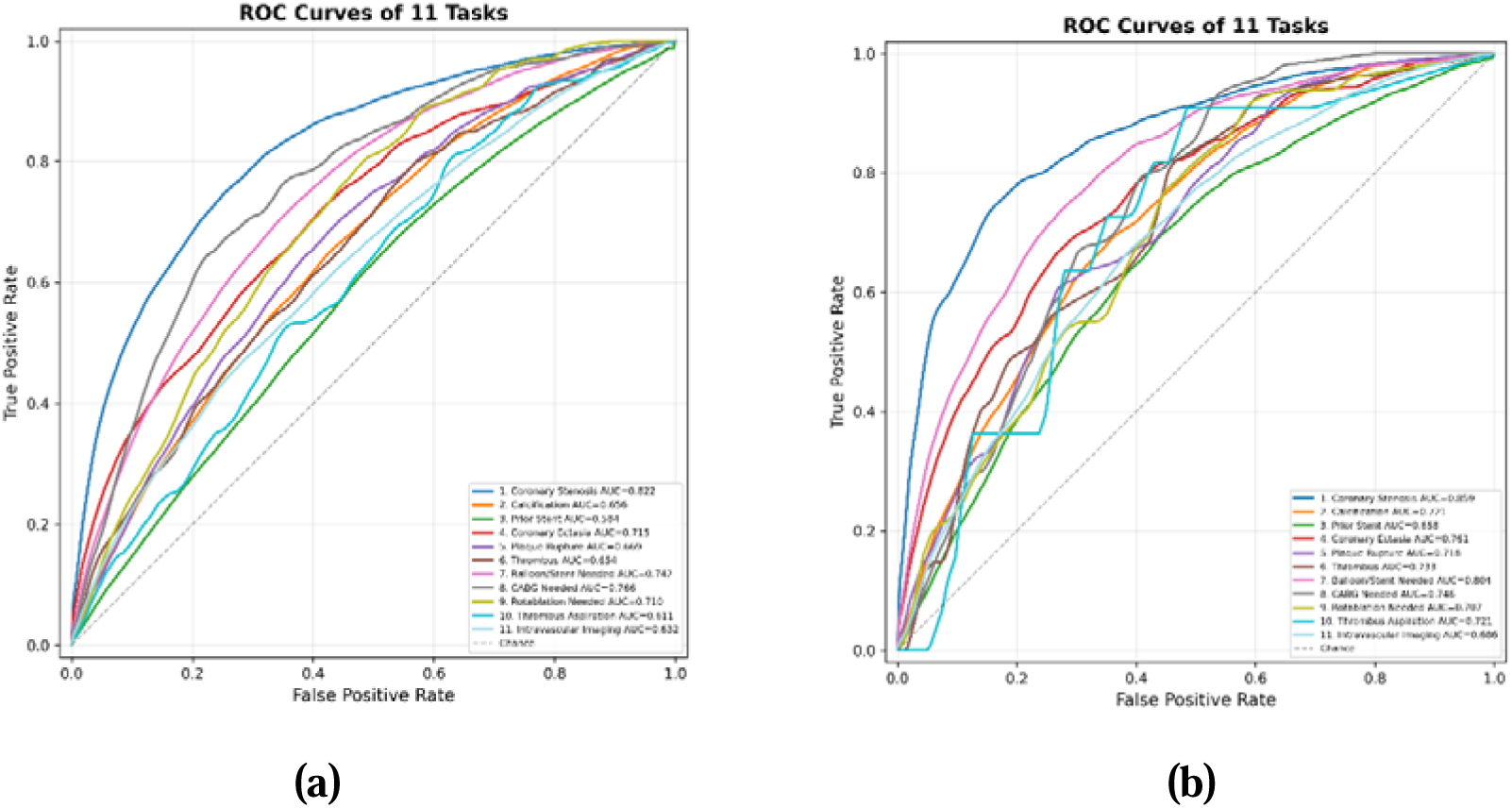
Zero-shot performance of CAG-MIND across multi-center coronary angiography tasks. (a) Receiver operating characteristic (ROC) curves of CAG-MIND for 11 downstream coronary angiography tasks evaluated in the Zhongshan Hospital internal validation cohort under the zero-shot setting. (b) ROC curves for the same tasks evaluated in the Shanghai Geriatric Medical Center external test cohort. The dashed diagonal line represents random classification (AUROC = 0.5). AUROC, area under the receiver operating characteristic curve; CABG, coronary artery bypass grafting.

**Table 2.** Zero-shot performance of CAG-MIND across multi-center coronary angiography tasks. The 11 downstream tasks were organized into three clinically relevant categories: **structural abnormality detection** (coronary stenosis, calcification, prior stent, and coronary ectasia), **atherosclerotic plaque assessment** (plaque rupture and thrombus), and **interventional decision prediction** (balloon/stent requirement, CABG recommendation, rotational atherectomy, thrombus aspiration, and intravascular imaging). Performance is reported as the area under the receiver operating characteristic curve (AUROC) with 95% confidence intervals. The final row shows the mean AUROC across all 11 tasks. CABG, coronary artery bypass grafting.

| Task category | Task | Zhongshan Hospital | Shanghai Geriatric Medical Center |
| --- | --- | --- | --- |
| Structural Abnormality Detection | Coronary Stenosis | 0.836 (0.826-0.845) | 0.864 (0.848-0.879) |
|  | Calcification | 0.652 (0.643-0.661) | 0.717 (0.698-0.737) |
|  | Prior Stent | 0.567 (0.558-0.574) | 0.688 (0.671-0.705) |
|  | Coronary Ectasia | 0.725 (0.707-0.743) | 0.752 (0.717-0.788) |
| Atherosclerotic Plaque Assessment | Plaque Rupture | 0.666 (0.647-0.686) | 0.716 (0.673-0.762) |
|  | Thrombus | 0.649 (0.609-0.686) | 0.759 (0.683-0.838) |
| Interventional Decision Prediction | Balloon/Stent Needed | 0.737 (0.730-0.743) | 0.814 (0.802-0.826) |
|  | CABG Needed | 0.749 (0.725-0.773) | 0.739 (0.694-0.782) |
|  | Rotablation Needed | 0.711 (0.683-0.739) | 0.715 (0.658-0.772) |
|  | Thrombus Aspiration | 0.620 (0.577-0.667) | 0.729 (0.618-0.832) |
|  | Intravascular Imaging | 0.636 (0.629-0.644) | 0.708 (0.692-0.726) |
| Average (11 tasks) |  | 0.686 | 0.745 |

CAG-MIND demonstrated robust zero-shot performance across all three task categories in both cohorts. Among structural abnormality detection tasks, coronary stenosis achieved the highest performance (AUROC 0.836 internally and 0.864 externally), while coronary ectasia also showed good discrimination. Plaque assessment tasks achieved moderate performance despite relying on more subtle angiographic features. Notably, the model maintained competitive performance for interventional decision prediction, achieving AUROCs of 0.737 and 0.814 for balloon/stent prediction and 0.749 and 0.739 for CABG prediction in the internal and external cohorts, respectively. Overall, the mean AUROC across all 11 tasks reached 0.686 in the internal validation cohort and 0.745 in the external test cohort, indicating that CAG-MIND learned transferable representations spanning coronary anatomy, plaque characteristics, and procedural decision-making without task-specific optimization.

### 3. Supervised fine-tuning performance across coronary angiography tasks

We next evaluated the performance of CAG-MIND after supervised fine-tuning across the same 11 clinically relevant coronary angiography tasks, organized into three hierarchical categories: structural abnormality detection, atherosclerotic plaque assessment, and interventional decision prediction, using the Zhongshan Hospital internal validation cohort and the Shanghai Geriatric Medical Center external test cohort (Fig. 5a,b and Table 3). Performance was quantified using the area under the receiver operating characteristic curve (AUROC) to assess task-specific adaptation following large-scale multimodal pretraining.

**Figure 5.**
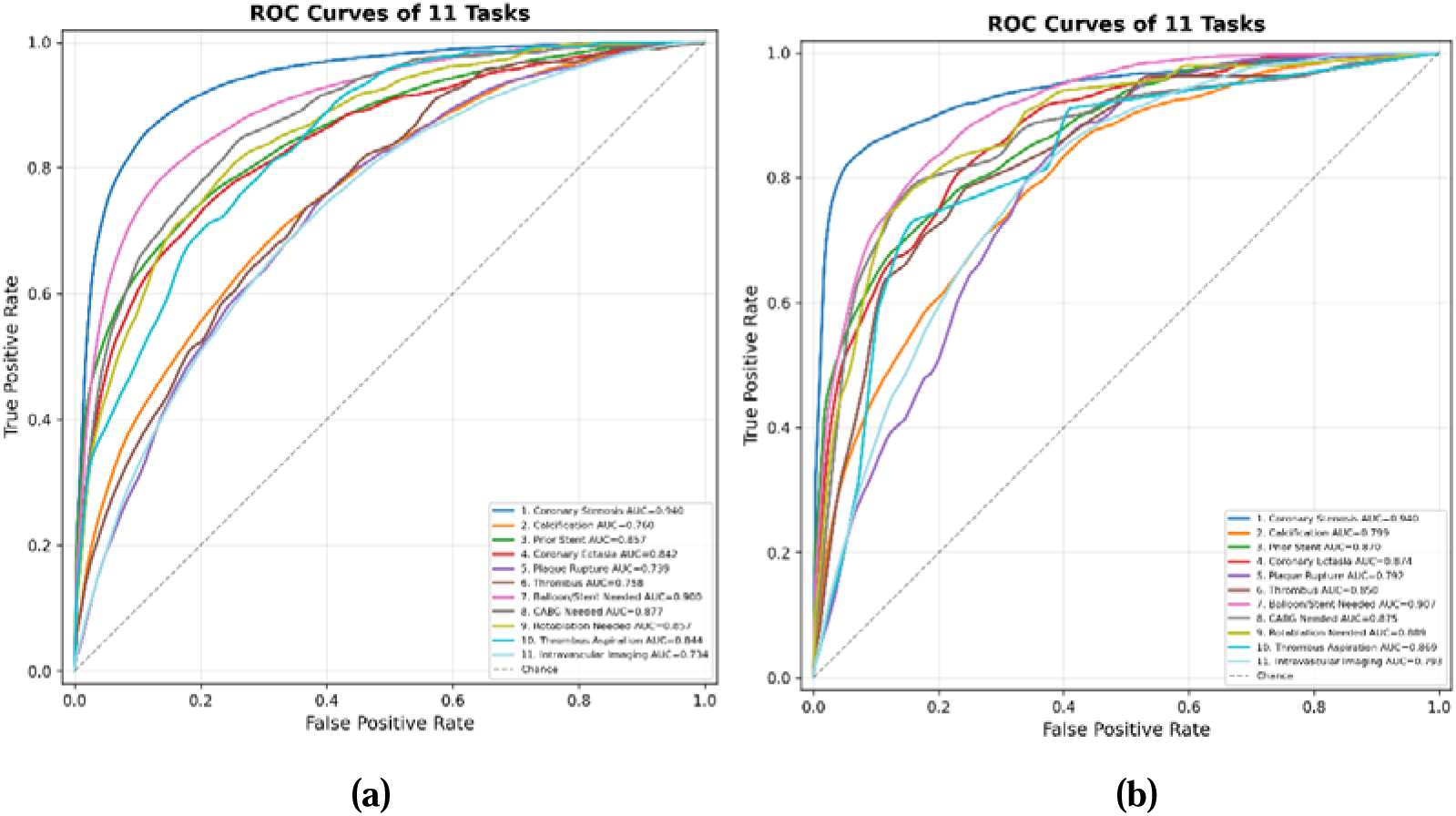
Supervised fine-tuning performance of CAG-MIND across multi-center coronary angiography tasks. (a) Receiver operating characteristic (ROC) curves of CAG-MIND after supervised fine-tuning for 11 downstream coronary angiography tasks in the Zhongshan Hospital internal validation cohort. (b) ROC curves for the same tasks in the Shanghai Geriatric Medical Center external test cohort. The dashed diagonal line represents random classification (AUROC = 0.5). AUROC, area under the receiver operating characteristic curve; CABG, coronary artery bypass grafting.

**Table 3.** Performance of CAG-MIND across three categories of coronary angiography tasks after supervised fine-tuning in the Zhongshan Hospital internal validation cohort and the Shanghai Geriatric Medical Center external test cohort. The 11 downstream tasks were grouped into three clinically relevant categories: structural abnormality detection (coronary stenosis, calcification, prior stent, and coronary ectasia), atherosclerotic plaque assessment (plaque rupture and thrombus), and interventional decision prediction (balloon/stent requirement, CABG recommendation, rotational atherectomy, thrombus aspiration, and intravascular imaging). Performance is reported as the area under the receiver operating characteristic curve (AUROC) with 95% confidence intervals. The final row presents the mean AUROC across all 11 tasks. CABG, coronary artery bypass grafting.

| Task category | Task | Zhongshan Hospital | Shanghai Geriatric Medical Center |
| --- | --- | --- | --- |
| Structural Abnormality Detection | Coronary Stenosis | 0.940 (0.935-0.945) | 0.940 (0.932-0.948) |
|  | Calcification | 0.760 (0.752-0.768) | 0.799 (0.783-0.815) |
|  | Prior Stent | 0.857 (0.852-0.862) | 0.870 (0.858-0.881) |
|  | Coronary Ectasia | 0.842 (0.828-0.856) | 0.874 (0.849-0.897) |
| Atherosclerotic Plaque Assessment | Plaque Rupture | 0.739 (0.721-0.756) | 0.792 (0.758-0.825) |
|  | Thrombus | 0.758 (0.725-0.789) | 0.850 (0.776-0.915) |
| Interventional Decision Prediction | Balloon/Stent Needed | 0.900 (0.896-0.904) | 0.907 (0.898-0.915) |
|  | CABG Needed | 0.877 (0.858-0.896) | 0.875 (0.823-0.921) |
|  | Rotablation Needed | 0.857 (0.835-0.879) | 0.889 (0.847-0.930) |
|  | Thrombus Aspiration | 0.844 (0.815-0.871) | 0.869 (0.782-0.941) |
|  | Intravascular Imaging | 0.734 (0.728-0.741) | 0.793 (0.779-0.806) |
| Average (11 tasks) |  | 0.827 | 0.846 |

Following supervised fine-tuning, CAG-MIND achieved consistently strong performance across all three task categories. In the internal validation cohort, structural abnormality detection demonstrated the highest overall performance, with AUROCs ranging from 0.760 to 0.940, led by coronary stenosis detection (AUROC, 0.940; 95% CI, 0.935–0.945). Plaque assessment tasks achieved AUROCs of 0.739 for plaque rupture and 0.758 for thrombus detection. For interventional decision prediction, the model demonstrated excellent discrimination for balloon/stent requirement (0.900) and CABG recommendation (0.877), while maintaining robust performance for rotational atherectomy (0.857), thrombus aspiration (0.844), and intravascular imaging prediction (0.734). The mean AUROC across all 11 tasks was 0.827.

Performance remained stable in the external test cohort, yielding a mean AUROC of 0.846 across all tasks. Coronary stenosis maintained excellent discrimination (AUROC, 0.940; 95% CI, 0.932–0.948), whereas plaque assessment and interventional decision prediction also showed consistently high performance, including thrombus detection (0.850), balloon/stent prediction (0.907), CABG recommendation (0.875), and rotational atherectomy prediction (0.889). Overall, the consistent performance observed across both centers demonstrates that large-scale multimodal pretraining enables effective supervised adaptation while preserving robust generalizability across diverse coronary angiography tasks.

### 4. Foundation-model performance compared with conventional architectures

We next compared the performance profiles of CAG-MIND with representative image-based and vision-language models across the 11 downstream coronary angiography tasks (Fig. 6a,b). For zero-shot evaluation, CAG-MIND was compared with BiomedCLIP and CLIP-CAG, whereas supervised fine-tuning performance was further compared with conventional image-based architectures, including 3DCNN, Xception, and Swin3D.

**Figure 6.**
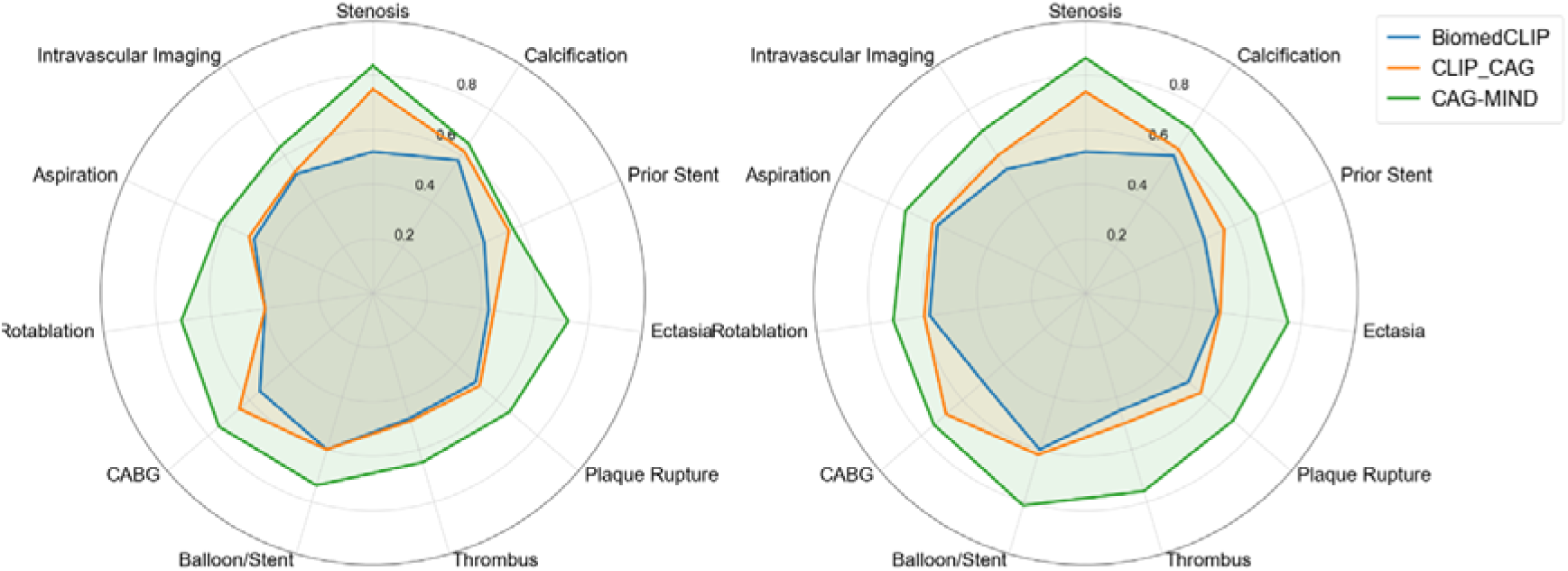
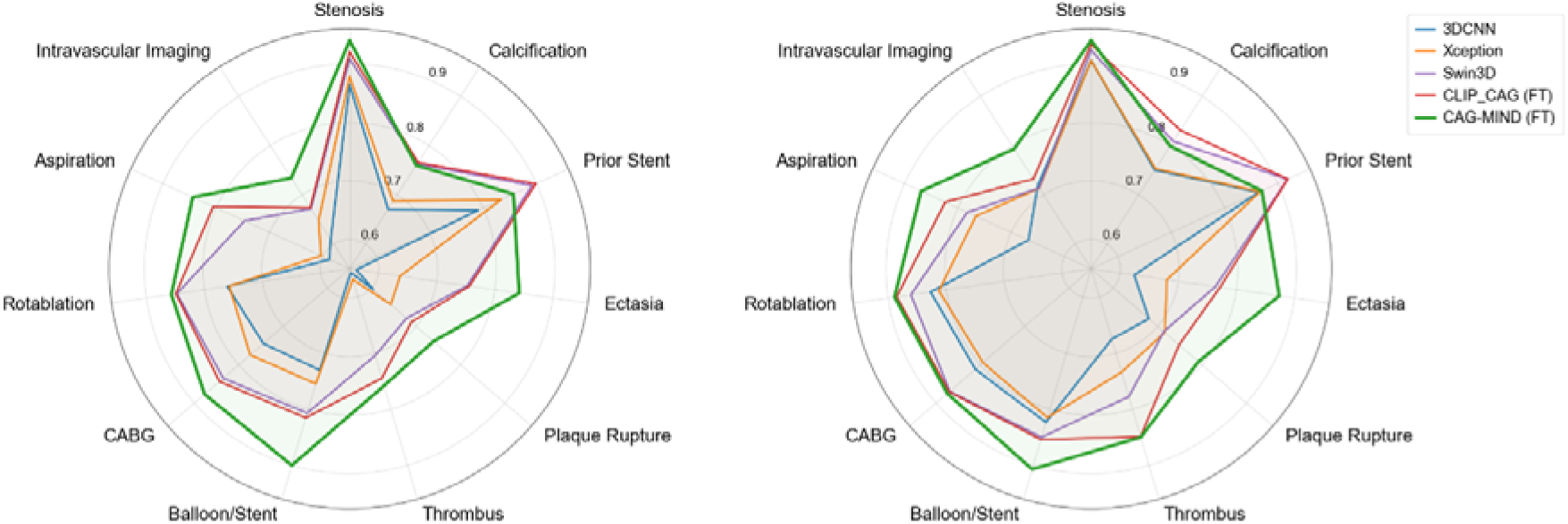
Comparison of task-specific performance profiles between CAG-MIND and representative image-based and vision-language models. **(a)** Radar plots showing zero-shot performance across 11 downstream coronary angiography tasks for BiomedCLIP, CLIP-CAG, and CAG-MIND in the Zhongshan Hospital internal validation cohort (left) and the Shanghai Geriatric Medical Center external test cohort (right). **(b)** Radar plots showing supervised fine-tuning performance for conventional image-based models (3DCNN, Xception, and Swin3D), the vision-language baseline (CLIP-CAG), and CAG-MIND in the corresponding internal and external cohorts. Higher values indicate better task-specific performance as measured by the area under the receiver operating characteristic curve (AUROC).

In the zero-shot setting, CAG-MIND consistently outperformed both BiomedCLIP and CLIP-CAG across all three task categories, including structural abnormality detection, atherosclerotic plaque assessment, and interventional decision prediction (Fig. 6a). The largest performance gains were observed for tasks requiring higher-level clinical understanding, such as coronary ectasia, plaque rupture, thrombus prediction, and interventional decision-making.

Following supervised fine-tuning, performance improved across all models; however, CAG-MIND maintained the best overall performance profile across nearly all downstream tasks (Fig. 6b). The preservation of this performance hierarchy suggests that the advantages conferred by large-scale domain-specific multimodal pretraining are retained after task-specific optimization rather than eliminated by supervised learning.

### 5. Model interpretability through Grad-CAM

Grad-CAM visualization was used to assess whether the model attends to clinically relevant regions in multi-view coronary angiograms. As shown in Fig. 7 (a,b), salient activations are predominantly localized to coronary vessel segments and suspected lesion-bearing regions, with limited attention allocated to surrounding background structures. This spatial concentration indicates that the model captures pathology-relevant imaging features rather than relying on nonspecific global appearance cues.

**Fig. 7.**
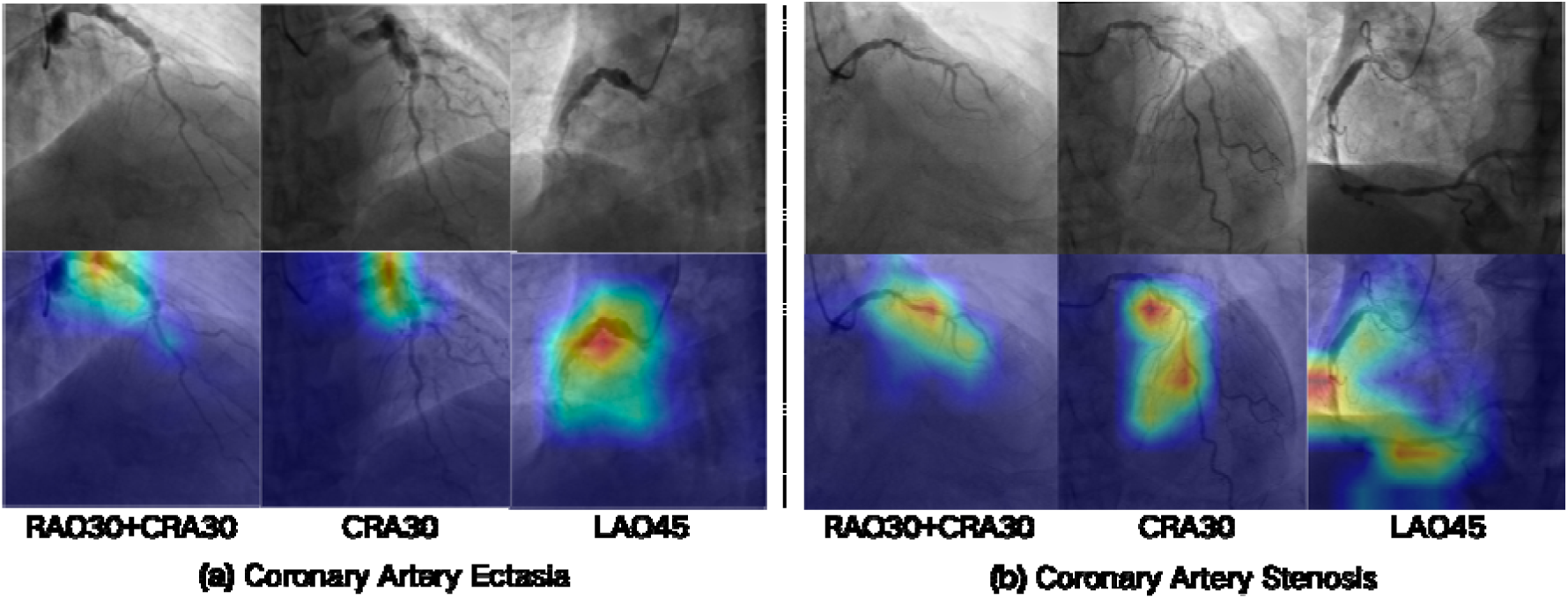
Grad-CAM visualization of spatial attention in multi-view coronary angiography. (a)Representative multi-view angiographic projections and corresponding Grad-CAM maps for coronary artery stenosis. (b) Representative multi-view angiographic projections and corresponding Grad-CAM maps for coronary artery ectasia. For both tasks, the top rows show original angiographic images and the bottom rows show the corresponding Grad-CAM maps. The activation regions are mainly localized along anatomically plausible coronary vessel segments and lesion-relevant areas, suggesting that the model focuses on task-relevant vascular morphology rather than irrelevant background structures. The view-dependent activation patterns further indicate adaptive use of complementary multi-view information across projections.

Importantly, the attention patterns differ across angiographic views, reflecting the view-dependent visibility of coronary anatomy and lesion morphology. In projections with clearer exposure of the vessel course, activations extend along the coronary lumen, whereas in other views the model focuses more narrowly on focal high-risk regions. These findings suggest that the network adaptively leverages complementary multi-view information and learns spatial representations that are aligned with clinically meaningful lesion localization.

Collectively, the spatial attention maps support the interpretability of the proposed framework by demonstrating that prediction-relevant features arise primarily from anatomically and pathologically plausible coronary regions.

## Discussion

### Principal findings

In this study, we developed CAG-MIND, a domain-specific vision-language foundation model for coronary angiography, and demonstrated that routine coronary angiography videos paired with procedural reports constitute a scalable multimodal substrate for foundation-model learning. Rather than optimizing individual downstream tasks, CAG-MIND learns a shared representation that integrates coronary anatomy, plaque characteristics, and procedural semantics from routine clinical data. Trained on more than 135,000 patients and over 800,000 angiographic videos, CAG-MIND represents, to our knowledge, the first coronary angiography vision-language model developed at true foundation-model scale.

Several findings support this concept. First, CAG-MIND consistently outperformed both general biomedical and coronary angiography-adapted vision-language baselines in zero-shot evaluation, indicating that domain-specific multimodal pretraining substantially improves representation quality for invasive cardiovascular imaging. Second, these advantages were preserved after supervised fine-tuning, suggesting that pretraining provides a stronger initialization rather than merely improving zero-shot inference. Third, CAG-MIND maintained competitive performance when fine-tuned using only 10% of labelled data, supporting data-efficient downstream adaptation. Finally, the model demonstrated stable external performance across institutions, indicating that the learned representations generalize beyond center-specific imaging characteristics.

Collectively, these findings suggest that CAG-MIND learns clinically meaningful representations extending beyond isolated visual pattern recognition toward integrated understanding of coronary anatomy, lesion morphology and procedural decision-making.

### Large-scale multimodal pretraining enables foundation-model learning for coronary angiography

Foundation models differ fundamentally from conventional supervised AI systems because their primary objective is to learn transferable representations rather than optimize individual downstream tasks. Recent multimodal medical foundation models such as BiomedCLIP, MedCLIP, CheXzero, EchoCLIP, and EchoPrime have demonstrated that aligning medical images with routinely generated clinical language provides scalable weak supervision capable of capturing clinically meaningful semantic representations without exhaustive manual annotation [17, 18, 20–22]. Our study extends this paradigm to invasive coronary angiography, a more challenging domain involving dynamic cine acquisition, multi-view dependency, vessel overlap, contrast dynamics, and procedural variability.

Importantly, procedural reports differ substantially from conventional image labels. Rather than describing isolated imaging findings, coronary angiography reports simultaneously encode anatomical distribution, lesion morphology, procedural interpretation and therapeutic decision-making. By aligning standardized six-view angiographic videos with these report-derived procedural semantics, CAG-MIND learns representations that reflect higher-level clinical concepts instead of individual visual attributes. We speculate that this richer semantic supervision contributes to the observed improvements in zero-shot transfer, data-efficient fine-tuning and external generalization.

The scale and standardization of the training dataset likely further contributed to these advantages. Large foundation models have consistently demonstrated that increasing data diversity and supervision scale improves robustness and transferability [23–26]. By combining more than 135,000 patients with standardized multi-view acquisition and report-derived weak supervision, CAG-MIND establishes a practical framework for foundation-model learning in invasive cardiovascular imaging.

### Relationship to previous coronary angiography AI studies

Prior AI studies in coronary angiography have mainly focused on isolated tasks, including stenosis grading, quantitative coronary analysis, angiography-derived physiological estimation, lesion segmentation, or plaque classification [9–11]. Computational physiology systems such as QFR and angiography-derived FFR have advanced non-wire-based hemodynamic assessment [27], while intravascular imaging modalities, including IVUS and OCT, have further improved plaque characterization and lesion assessment [28,29].

In contrast, CAG-MIND adopts a unified multimodal foundation-model framework that simultaneously encodes anatomical findings, plaque characteristics, and procedural semantics within a shared representational space. Its zero-shot transfer capability further distinguishes it from conventional supervised models. This is important because coronary angiography interpretation rarely depends on a single visual feature, but instead requires integration of anatomy, lesion morphology, vessel distribution, and procedural feasibility.

### Scientific interpretation of task-specific performance differences

A central finding was the non-uniform distribution of performance gains across tasks. Morphology-dominant tasks, such as stenosis detection, calcification assessment, and prior stent identification, showed consistent but relatively moderate improvements. These tasks rely mainly on direct angiographic features, including luminal narrowing, radiopacity, vessel contour, and stent-like density. Conventional image-based models can therefore capture part of the relevant signal after supervised training.

In contrast, larger gains were observed in lesion-context and procedural prediction tasks, including plaque rupture, thrombus, rotational atherectomy prediction, CABG referral, balloon/stent requirement, thrombus aspiration, and intravascular imaging use. These tasks require integration of lesion location, vessel size, lesion length, bifurcation involvement, calcification burden, multivessel distribution, and procedural feasibility. They are therefore more likely to benefit from multimodal representation learning.

The improved performance in interventional decision prediction suggests that vision-language alignment may encode latent associations between angiographic phenotypes and procedural intent derived from routine reports. However, this should not be interpreted as proof of true clinical reasoning. Rather, CAG-MIND appears to learn representations more closely aligned with clinically contextualized procedural patterns than isolated visual classifiers.

### Cross-center robustness and generalizability

The stable performance of CAG-MIND across independent centers supports the hypothesis that multimodal pretraining improves domain generalization. Distributional shift remains a major barrier to clinical AI deployment, as many deep learning systems degrade under external validation because of shortcut learning and center-specific bias [30–32]. The preserved performance hierarchy across cohorts suggests that domain-aligned multimodal supervision may reduce reliance on acquisition-specific correlations and promote learning of disease-relevant imaging features.

This is particularly relevant for coronary angiography, where image appearance varies with imaging systems, operator technique, catheter engagement, contrast injection, projection angle, frame rate, and institutional protocols. Grad-CAM visualizations further showed that the model primarily attended to anatomically plausible coronary vessel regions and lesion-relevant areas, with view-dependent activation patterns suggesting adaptive use of complementary projections. Nevertheless, broader validation across additional populations, imaging systems, and healthcare environments remains necessary.

### Modality-specific considerations

Coronary angiography has intrinsic modality-specific constraints. Conventional luminography incompletely characterizes plaque composition, vessel wall biology, microvascular dysfunction, and dynamic hemodynamic significance [5–7,36]. Thus, some pathological states may remain difficult to infer from angiography alone, even with advanced representation learning.

This limitation may explain part of the task-level variability. Stenosis, stent presence, and severe calcification are relatively direct angiographic findings, whereas plaque rupture, thrombus, microvascular dysfunction, and physiological significance may be only indirectly reflected by angiographic appearance. Future models may therefore benefit from integration with IVUS, OCT, physiological assessment, and longitudinal clinical data.

Although Grad-CAM showed anatomically plausible attention, explainability methods remain imperfect and should be interpreted cautiously [37,38]. Attention localization does not prove causal reasoning or guarantee clinically interpretable decision processes. Further prospective validation, counterfactual testing, failure-mode analysis, and physician-AI interaction studies are needed.

### Clinical implications

CAG-MIND may support automated or semi-automated coronary angiography interpretation by identifying stenosis, calcification, prior stents, ectasia, thrombus, and other lesion-related features. This may reduce inter-operator variability and improve reporting consistency, particularly in high-volume catheterization laboratories.

The model may also assist procedural decision support. Its performance in predicting balloon/stent requirement, CABG recommendation, rotational atherectomy, thrombus aspiration, and intravascular imaging use suggests potential value in providing preliminary procedural suggestions or highlighting cases requiring expert review. Such use may be helpful for junior operators, heart-team discussions, and institutions with variable interventional expertise.

In addition, its data-efficient fine-tuning performance suggests that CAG-MIND could serve as a foundation model for downstream tasks, such as culprit lesion identification, SYNTAX score estimation, no-reflow risk prediction, PCI complexity assessment, LVEF prediction from angiography, or long-term outcome prediction. Future cath-lab intelligence systems may further integrate angiography with IVUS, OCT, FFR/iFR/QFR, ECG, echocardiography, laboratory tests, medications, devices, and longitudinal outcomes [33–36].

These applications should be positioned as decision support rather than replacement of physician judgement. Final diagnosis and treatment decisions must remain under expert clinical supervision.

### Limitations

Several limitations should be acknowledged. First, this was a retrospective study, and prospective catheterization laboratory studies are required to assess real-time utility, workflow feasibility, and impact on clinical decision-making. Second, although external validation was performed, broader multinational and multi-vendor validation is necessary. Third, supervision was derived from routine procedural reports, which may contain operator-dependent bias, institutional practice patterns, incomplete descriptions, and terminology variability. Fourth, Grad-CAM provides only approximate visualization and does not prove causal reasoning or guarantee interpretability. Finally, whether CAG-MIND improves patient outcomes, reduces procedure time, improves treatment selection, or lowers complication rates remains unknown and requires prospective clinical trials or pragmatic implementation studies.

## Conclusion

In conclusion, CAG-MIND establishes a scalable vision-language foundation-model paradigm for coronary angiography and invasive cardiovascular imaging. By aligning standardized multi-view angiographic videos with structured procedural semantics at unprecedented scale, it enables robust zero-shot transfer, data-efficient fine-tuning, and cross-center generalization across diverse diagnostic and interventional tasks. The structured pattern of task-level improvements suggests that domain-specific multimodal pretraining captures clinically meaningful associations between coronary anatomy, lesion morphology, and procedural decision-making. With further prospective validation and multimodal integration, CAG-MIND may serve as a foundation for future intelligent decision-support systems in interventional cardiology.

## Data Availability

All data produced in the present study are available upon reasonable request to the authors。

## Supplement figure

**Fig. S1.**
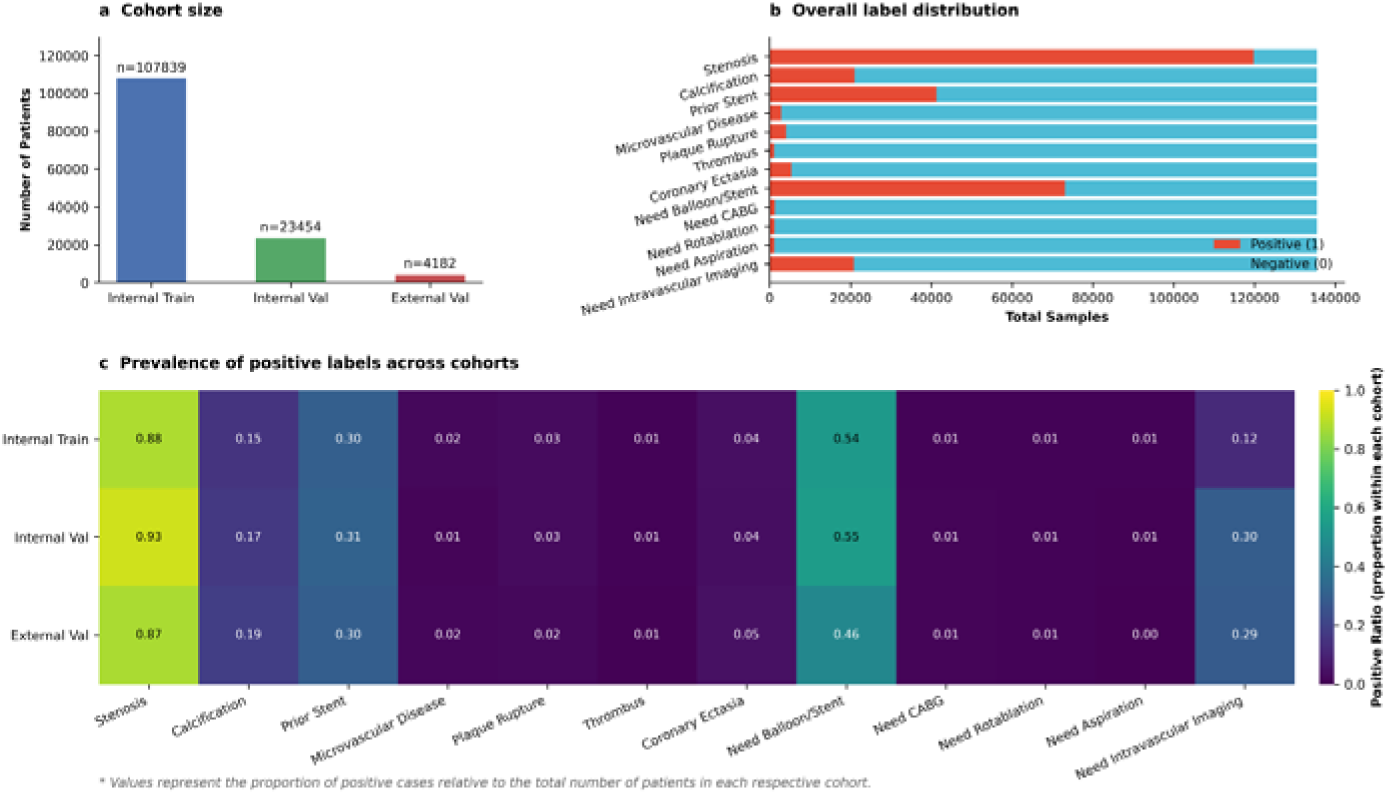
Cohort size and label distribution of the CAG-MIND dataset. The dataset comprises 107,839 internal training cases, 23,454 internal validation cases and 4,182 external validation cases. Overall label frequencies show substantial variation across tasks, and the prevalence of positive labels is largely consistent across cohorts.

## Reference

1. Lawton JS, Tamis-Holland JE, Bangalore S, et al. 2021 ACC/AHA/SCAI Guideline for Coronary Artery Revascularization. J Am Coll Cardiol. 2022;79:e21–e129. doi:10.1016/j.jacc.2021.09.006

2. Patel MR, Dehmer GJ, Hirshfeld JW, et al. ACCF/SCAI/AATS/AHA/ASE/ASNC/HFSA/HRS/SCAI/SCCT/SCMR Appropriate Use Criteria for Diagnostic Catheterization. J Am Coll Cardiol. 2012;59:1995–2027. doi:10.1016/j.jacc.2012.03.003

3. Dey D, Slomka PJ, Leeson P, et al. Artificial Intelligence in Cardiovascular Imaging: JACC State-of-the-Art Review. J Am Coll Cardiol. 2019;73:1317–1335. doi:10.1016/j.jacc.2018.12.054

4. Topol EJ. High-performance medicine: the convergence of human and artificial intelligence. Nat Med. 2019;25:44–56. doi:10.1038/s41591-018-0300-7

5. Ouyang D, He B, Ghorbani A, et al. Video-based AI for beat-to-beat assessment of cardiac function. Nature. 2020;580:252–256. doi:10.1038/s41586-020-2145-8

6. Zhang J, Gajjala S, Agrawal P, et al. Fully automated echocardiogram interpretation in clinical practice. Circulation. 2018;138:1623–1635. doi:10.1161/CIRCULATIONAHA.118.034338

7. Attia ZI, Kapa S, Lopez-Jimenez F, et al. Screening for cardiac contractile dysfunction using an artificial intelligence-enabled electrocardiogram. Nat Med. 2019;25:70–74. doi:10.1038/s41591-018-0240-2

8. Hannun AY, Rajpurkar P, Haghpanahi M, et al. Cardiologist-level arrhythmia detection with convolutional neural networks. Nat Med. 2019;25:65–69. doi:10.1038/s41591-018-0268-3

9. Itu L, Rapaka S, Passerini T, et al. A machine-learning approach for computation of fractional flow reserve from coronary computed tomography. J Appl Physiol. 2016;121:42–52. doi:10.1152/japplphysiol.00752.2015

10. Zreik M, Lessmann N, van Hamersvelt RW, et al. Deep learning analysis of coronary arteries in cardiac CT angiography for detection of patients requiring revascularization. JACC Cardiovasc Imaging. 2020;13:1545–1557. doi:10.1016/j.jcmg.2019.07.030

11. Bernardo, R., Nurmohamed, N. S., Bom, M. J. et al Diagnostic accuracy in coronary CT angiography analysis: artificial intelligence versus human assessment. Open heart, 2025 12(1), e003115. 10.1136/openhrt-2024-003115

12. Johnson KW, Torres Soto J, Glicksberg BS, et al. Artificial intelligence in cardiology. J Am Coll Cardiol. 2018;71:2668–2679. doi:10.1016/j.jacc.2018.03.521

13. Radford A, Kim JW, Hallacy C, et al. Learning transferable visual models from natural language supervision. Proc Mach Learn Res. 2021;139:8748–8763.

14. Jia C, Yang Y, Xia Y, et al. Scaling up visual and vision-language representation learning with noisy text supervision. Proc Mach Learn Res. 2021;139:4904–4916.

15. Li J, Li D, Savarese S, et al. BLIP-2: Bootstrapping language-image pre-training with frozen image encoders and large language models. arXiv. 2023. doi:10.48550/arXiv.2301.12597

16. Bommasani R, Hudson DA, Adeli E, et al. On the opportunities and risks of foundation models. arXiv. 2021. doi:10.48550/arXiv.2108.07258

17. Christensen M, Vukadinovic M, Yuan N, et al. Vision-language foundation model for echocardiogram interpretation. Nat Med. 2024;30:1481–1488. doi:10.1038/s41591-024-02959-y

18. Vukadinovic M, Tang X, Yuan N, et al. EchoPrime: A multi-video view-informed vision-language model for comprehensive echocardiography interpretation. arXiv. 2024. doi:10.48550/arXiv.2410.09704

19. Ahmed, Imtiaz, et al. “Qwen 2.5: A comprehensive review of the leading resource-efficient llm with potentioal to surpass all competitors.” techrXiv, 2025. 10.36227/techrxiv.174060306.65738406/v1

20. Wang Z, Liu Y, Wang H, et al. MedCLIP: contrastive learning from unpaired medical images and text. EMNLP. 2022.

21. Zhang S, Xu H, Usuyama N, et al. BiomedCLIP: a multimodal biomedical foundation model pretrained from fifteen million scientific image-text pairs. arXiv. 2023. doi:10.48550/arXiv.2303.00915

22. Tiu E, Tali D, Shah P, et al. Expert-level detection of pathologies from unannotated chest X-ray images via self-supervised learning. Nat Biomed Eng. 2022;6:1399–1406. doi.org/10.1038/s41551-022-00936-9

23. Moor M, Banerjee O, Abad ZSH, et al. Foundation models for generalist medical artificial intelligence. Nature. 2023;616:259–265. doi:10.1038/s41586-023-05881-4

24. Singhal K, Tu T, Gottweis J, et al. Towards expert-level medical question answering with large language models. Nature medicine, 31(3), 943–950.doi.org/10.1038/s41591-024-03423-7.

25. Kelly, C. J., Karthikesalingam, A., Suleyman, M., Corrado, G., & King, D.. Key challenges for delivering clinical impact with artificial intelligence. BMC medicine, 2019, 17(1), 195. doi.org/10.1186/s12916-019-1426-2

26. Fearon WF, Nishi T, De Bruyne B, et al. Clinical outcomes and cost-effectiveness of fractional flow reserve-guided percutaneous coronary intervention in patients with stable coronary artery disease. Circulation. 2018;137:480–487. doi:10.1161/CIRCULATIONAHA.117.031907

27. Tonino PAL, De Bruyne B, Pijls NHJ, et al. Fractional flow reserve versus angiography for guiding percutaneous coronary intervention. N Engl J Med. 2009;360:213–224. doi:10.1056/NEJMoa0807611

28. Nissen SE, Yock P. Intravascular ultrasound: novel pathophysiological insights and current clinical applications. Circulation. 2001;103:604–616. doi:10.1161/01.CIR.103.4.604

29. Prati F, Guagliumi G, Mintz GS, et al. Expert review document on methodology, terminology, and clinical applications of optical coherence tomography. Eur Heart J. 2010;31:401–415. doi:10.1093/eurheartj/ehp433

30. Selvaraju RR, Cogswell M, Das A, et al. Grad-CAM: visual explanations from deep networks via gradient-based localization. Proc IEEE Int Conf Comput Vis. 2017:618–626. doi:10.1109/ICCV.2017.74

31. Jain S, Wallace BC. Attention is not explanation. NAACL. 2019:3543–3556. doi:10.18653/v1/N19-1357

32. Krittanawong C, Johnson KW, Rosenson RS, et al. Deep learning for cardiovascular medicine: a practical primer. Eur Heart J. 2019;40:2058–2073. doi:10.1093/eurheartj/ehz056

33. Tu S, Westra J, Yang J, et al. Diagnostic accuracy of fast computational approaches to derive fractional flow reserve from diagnostic coronary angiography. JACC Cardiovasc Interv. 2016;9:2024–2035. doi:10.1016/j.jcin.2016.07.013

34. Morris PD, Ryan D, Morton AC, et al. Virtual fractional flow reserve from coronary angiography: modeling the significance of coronary lesions. JACC Cardiovasc Interv. 2013;6:149–157. doi:10.1016/j.jcin.2012.08.024

35. Fearon WF, Achenbach S, Engstrom T, et al. Accuracy of fractional flow reserve derived from coronary angiography. Circulation. 2019;139:477–484. doi.org/10.1161/CIRCULATIONAHA.118.037350

36. Kany, S., Friedman, S. F., Al-Alusi, M., et al. Electrocardiogram-Based Artificial Intelligence to Identify Coronary Artery Disease. JACC. Advances, 2025 4(9), 102041. 10.1016/j.jacadv.2025.102041

